# DERIVED-BAND AUDITORY BRAINSTEM RESPONSES: COCHLEAR CONTRIBUTIONS DETERMINED BY NARROWBAND MASKERS

**DOI:** 10.1101/2023.05.13.23289939

**Authors:** David R. Stapells, Maxine R. Fok

## Abstract

**Objective:** The present study sought to determine the cochlear frequency regions represented by Audtory Brainstem Responses (ABRs) obtained using the high-pass noise/derived response (HP/DR) technique.

**Design:** Broadband noise sufficient to mask the ABR to 50 dB nHL clicks was HP filtered (96 dB/oct) at 8000, 4000, 2000, 1000 and 500 Hz. Mixed with the clicks and high-pass noise masker was narrowband noise. Three DR bands, denoted by the upper and lower high-pass noise frequencies, were obtained: DR4000-2000, DR2000-1000, and DR1000-500.

**Study sample:** Ten adults with normal hearing, aged 19-27 years (mean age: 22.4 years), were recruited from the community.

**Results:** Frequencies contributing to each DR were determined from the wave V percent amplitude (or latency shift) vs narrowband masker frequency profiles (relative to a no-narrowband-noise condition). Overall, results indicate derived band center frequencies were closer to the lower HP cutoff frequencies for DR4000-2000 and DR2000-1000, and approximately halfway between the lower HP cutoff and the geometric mean of the two HP frequencies for DR1000-500, with bandwidths of 0.5-1 octave in width.

**Conclusions:** These results confirm the validity of the HP/DR technique for assessing narrow cochlear regions (≤1.0 octave wide), with center frequencies within ½-octave of the lower HP cutoff frequency.

## Introduction

The relative contribution of specific portions of the basilar membrane to a surface-recorded response, such as the auditory brainstem response (ABR), may be revealed by the “high-pass noise/derived response (HP/DR)” technique. The HP/DR technique was originally introduced for recordings of cochlear nerve compound action potentials in animals by Teas and colleagues (Teas, Eldridge, and Davis 1962), and subsequently applied to human electrocochleographic recordings by Elberling (1974) and by Eggermont (1976) and soon after to the ABR by Don and Eggermont (1978) as well as by Parker and Thornton (1978b). Since these first reports, the HP/DR technique has proven to be an extremely valuable tool for investigation of mechanisms underlying the ABR, and, to a lesser extent, for clinical investigations. It has been used with success for many purposes, including: evaluation of the cochlear contributions to the auditory evoked potential to a specific acoustic stimulus (“cochlear place specificity”) (Burkard and Hecox 1983; Burkard and Voigt 1989; Don and Eggermont 1978; Elberling 1974; Herdman, Picton, and Stapells 2002; Kramer 1992; Laukli and Mair 1986; Nousak and Stapells 1992; Oates and Stapells 1997; Parker and Thornton 1978b), estimation of threshold for specific frequencies (e.g., Don, Eggermont, and Brackmann 1979; Koide, Yoshie, and Ase 1985), determination of travelling wave velocity, such as in Ménière’s patients (e.g., Parker and Thornton 1978a; Thornton and Farrell 1991), investigation of ABR sex differences (e.g., de Boer, Hardy, and Krumbholz 2022; Don et al. 1993; Ponton et al. 1992), investigation of ABR changes with maturation (e.g., Ponton et al. 1992), and improvement in acoustic tumor detection by ABR (e.g., Don et al. 1997). More recently, results from ABR derived response latencies have also been used for the design and analysis of chirp stimuli (de Boer, Hardy, and Krumbholz 2022; Elberling and Don 2008; Wegner and Dau 2002).

In the HP/DR technique, the ABR to a stimulus is recorded in quiet and then simultaneously with broadband noise of sufficient intensity to completely mask the ABR. This broadband noise is then steeply high-pass (HP) filtered, and the ABR then recorded in the presence of the HP noise masking. The response recorded in HP noise at one cutoff frequency is subtracted from the response obtained in the presence of HP noise with a higher cutoff frequency. The result is a response reflecting a “derived band” of frequencies approximately between the two cutoff frequencies. However, due to the finite slope of the HP filter, some masking below the lowest HP cutoff frequency occurs, and the derived response resulting from the subtraction of the two HP noise-masked responses will therefore include frequencies below, as well as above, the lowest HP frequency (Don and Eggermont 1978; Eggermont and Don 1980; Oates 1996). Separations of one octave of HP noise cutoffs are typically employed.

The validity of the HP/DR technique has been proven in several ways. Using electrocochleography and studying guinea pigs, Teas and colleagues (Teas, Eldridge, and Davis 1962) chemically inactivated relatively large portions of the cochlear partition (basal end vs apical 2/3rds), and demonstrated reduction of responses in DR bands representing these regions. Evans and Elberling (1982) recorded responses to click stimuli in the presence of HP noise from 27 individual cochlear nerve fibers of a cat, and concluded assumptions about HP noise masking were substantiated, at least for frequencies above 2 kHz (and likely for 0.5-1kHz). Parker and Thornton (1978c) recorded DR ABRs in humans and, assessing the effects of three different maskers (#1: <750Hz; #2: 1010-3000 Hz; #3: >4040 Hz), determined that if the masker frequencies were between the HP cutoffs, the resulting DR ABR amplitudes/presence were reduced. Several studies assessed humans with hearing loss, comparing pure-tone behavioral thresholds with those of DR bands, using 1-octave-wide bands and steps. These studies showed elevated thresholds for the DR ABR bands corresponding to the elevated behavioral thresholds (e.g., Don, Eggermont, and Brackmann 1979; Koide, Yoshie, and Ase 1985).

There is some disagreement concerning the center frequency (CF) of a derived band. Some studies have estimated the CF by subtracting the spectrum of the HP noise at one cutoff frequency from the spectrum of the HP noise at the higher cutoff frequency, and then taking the centre of this estimated derived band. Using this method, the CF is typically close to the lower HP cutoff frequency for one-octave-wide separations (and below the lower HP cutoff frequency for half-octave-wide separations), thus many studies have used the lower HP cutoff frequency as the derived-band CF (e.g., Don, Eggermont, and Brackmann 1979; Eggermont and Don 1980; Herdman, Picton, and Stapells 2002; Kramer 1992; Nousak and Stapells 1992; Oates and Stapells 1997). Most later studies (including, since about 1992, those of Don and colleagues), however, have calculated the geometric mean of the two HP cutoffs, and used this as the CF (e.g., de Boer, Hardy, and Krumbholz 2022; Don et al. 1993; Don et al. 1997; Elberling and Don 2008; Finneran et al. 2016). Both of the above methods are arbitrary: the use of the geometric mean does not take into account characteristics of the HP noise, such as its slope (indeed, use of the geometric mean assumes a slope of infinite steepness). Neither the geometric mean nor the CF calculation from the HP noise spectra take into account cochlear, eighth nerve, or brainstem physiology (including, cochlear upward spread of excitation, cochlear/eighth nerve two-tone suppression, or brainstem interactions). Some studies avoid the issue by simply naming a DR band by the two HP noise cutoffs (such as: DR2000-1000) (e.g., Burkard and Voigt 1989; Elberling 1974; Evans and Elberling 1982; Koide, Yoshie, and Ase 1985; Parker and Thornton 1978a).

Although the ABR HP/DR technique (or data from the technique) has been used for over 40 years and in more than 50 studies, there do not appear to be any studies investigating in detail the cochlear contributions to the “derived band” ABR. None of the studies described above were designed to determine the center frequencies or bandwidths of any DR band. In the present study, we sought to directly determine the center frequencies and bandwidths of the HP/DR band ABRs by using narrowband maskers within the high-pass noise, and thus within the derived bands.

## Methods

### Subjects

Ten adults (5 males) with normal hearing, aged 19-27 years (mean: 22.4 years), were paid to participate in this study. Normal hearing, for this study, was defined as pure-tone behavioral thresholds in the test ear of 15 dB HL (ANSI 1996) or better for each of 500, 1000, 2000 and 4000 Hz. To ensure results across the derived bands would not be influenced by differences in hearing across frequency, an additional criterion stipulated that there be no more than a 10-dB difference between any of these frequencies. Mean thresholds were 7.0, 4.0, 3.0, 2.0, −0.5 and 4.5 dB HL for 250, 500, 1000, 2000, 4000 and 8000 Hz, respectively. On each day of testing, subjects had normal tympanograms and present ipsilateral acoustic reflexes in their test ear.

### Stimuli

The stimuli were clicks (60-µs square wave) presented via an ER-3A insert earphone at 50 dB nHL (74.6 dB ppe SPL, DB0138 2-cc coupler, Quest model 1800 sound level meter), using a 59/s stimulus rate (Neuroscan STIM, digital-to-analog rate of 60,000 Hz). The acoustic spectrum of the 60-µs square wave presented through the ER-3A earphone was reasonably flat from <100Hz up to about 4000 Hz, then decreasing to −20, −30 and −40 dB at 5000, 6000 and 7000 Hz, respectively. Assuming an effective bandwidth of about 5000 Hz, the level per Hz (No) of these 50 dB nHL clicks is about 38 dB SPL.

### High-pass (HP) noise maskers

Broadband masking noise (TDT WG-1, 20kHz bandwidth, white noise) was high-pass filtered (Wavetek 852, 96 dB/octave slope), attenuated, and mixed with the click stimuli (and BBN masker, see below). This is the same setup (and equipment) as used in our previous research (Herdman, Picton, and Stapells 2002). Recordings were obtained in Quiet (no noise), broadband masking (to determine level required for HP noise), and HP noise with cutoffs of 8000, 4000, 2000, 1000, and 500 Hz. Within each high-pass noise condition (except 8000 Hz), there were nine NBN masker sub-conditions plus one no-narrowband-masker sub-condition, giving 10 sub-conditions.

The level of the HP noise was determined as follows: (i) the level of broadband masker (Wavetek filter set to HP 0.1 Hz) required to just behaviorally mask the 50 dB nHL clicks was determined for each subject using a 2-dB-down/1-dB-up bracketing procedure; (ii) the masking level was increased to 7 dB above the behavioral masking level to ensure the ABR would be masked; and (iii) recordings (2 replications) were then obtained in this broadband masking noise to ensure the ABR was completely masked. If it was not masked, recordings at higher noise levels (1-dB steps) were obtained until no response was present. Mean masking levels for the 10 subjects were 70.4 (behavioral; range: 67.5-74.5) and 82.0 (ABR; range: 77.5-87.5) dB SPL.

### Narrowband noise (NBN) maskers

1/3rd-octave-wide NBN maskers were used to assess the bandwidths and center frequencies of the derived bands. White noise (General Radio 1381, 2Hz-50kHz) was bandpass filtered using two cascaded bandpass filters: SRS SR650 (115 dB/octave slope), cascaded to a TDT PF1 filter (>1kHz: 60 dB/octave slope; <1kHz: 36 dB/octave slope), both set with a 1/3-octave-wide passband. The resulting maskers show a narrowband (1/3 octave) of energy in the passband and levels outside the passband that are at least −50 dB (at ½-octave above and below the passband) relative to the passband. NBN maskers had center frequencies from 250 Hz to 6350 Hz (using 1/3-octave steps). Each NBN masker was mixed with the click stimuli and high-pass noise masker (TDT SM3). The level of each NBN masker was set so that the level per cycle (No) of the masker was 43 dB SPL, about 5 dB higher than that of the click level per cycle.^1^

### ABR Recordings

All recordings and waveform analyses were carried out using a Neuroscan SYNAMPS/SCAN system. Recordings were obtained using electrodes placed on the vertex (Cz, noninverting), ipsilateral earlobe (inverting), and C7 vertebra (ground). Interelectrode impedances were 3000 Ohms or less. The electroencephalographic (EEG) signals were amplified (SYNAMPS) and, using Neursocan SCAN software, single-trial epochs over a 5-20 ms post-stimulus analysis time digitized (100,000 Hz sampling rate) and digitally filtered (30-1500 Hz, 12 dB/octave). Trials with amplitudes exceeding ±200 µV were automatically rejected. The combination of the 59/s stimulus rate and beginning the analysis period 5 ms following the click allowed a faster rate (resulting in more-reasonable test times) *and* ensured wave V, when present, would be captured for all conditions.

Single-trial epochs were averaged using the Neuroscan SCAN software online Fsp/Bayesian averaging software, which allows one to (i) terminate the recording when a specified estimated residual noise level (RNL) is attained, and (ii) weights blocks of trials (256 trials each) based on their RNL. In the present study, data collection for each replication was terminated when the single-point (at 12.5 ms post stimulus) estimate of RNL had decreased to 40 nanovolts. Three replications were obtained in each masked condition.

### Derived-band waveforms

Derived brainstem responses (DR) were obtained by successive subtraction of a waveform recorded in one high-pass noise from the waveform recorded in high-pass noise with a 1-octave higher cutoff. Thus, the following bands were derived and designated as: DR4000-2000 (i.e., HP4000 minus HP2000), DR2000-1000, and DR1000-500. Within each HP noise condition were 9 narrowband masker sub-conditions (plus one without NBN). Each derived-response subtraction was carried-out for waveforms within the *same* NBN sub-condition. Thus, in addition to a no-NBN sub-condition for each derived band, we obtained DR4000-2000 results for narrowband maskers (1/3-octave steps) with CFs from 1000 to 6350 Hz, DR2000-1000 results for narrowband maskers with CFs from 500 to 3175 Hz, and DR1000-500 results for narrowband maskers with CFs from 250 to 1587 Hz. As we also recorded the ABRs in HP8000 noise, we also calculated the DR8000-4000 DRs; the effects of NBN maskers, however, were not assessed for this DR.

The derived responses were calculated as described above for each of three replications obtained in each sub-condition. The subtraction procedure theoretically increased the estimated RNL of each replication to 56.6 nanovolts. An overall average of the three DR replications would theoretically have an estimated RNL of 32.7 nanovolts, allowing peaks as small as 0.1-0.2 µV to be reliably measured (Don and Elberling 1996).

### Response measures

Waveforms (3 replications and overall average for each sub-condition) were assessed by two individuals familiar with ABR recordings and their measurement. Response presence required a repeatable peak with a peak-to-peak amplitude of at least 0.1 µV. Wave measures (wave V latency, corrected for insert earphone and trigger delay; V-V’ peak-to-peak amplitude) were made from the average of the replications. Wave V’ was defined as the greatest negativity occurring within 8 ms after wave V (e.g., Nousak and Stapells 1992; Oates and Stapells 1997). If no response was present, the estimated RNL was entered as the amplitude.^2^

Subjects’ latencies and amplitudes in each NBN sub-condition were normalized relative to their results for the appropriate no-NBN (“non-masked”) sub-condition for the same derived band. Plotting these results as a function of narrowband CF provides latency-shift and percent-of-non-masked-amplitude profiles for each derived band. Estimates of the CF and bandwidth of the three derived bands were obtained from each subject’s amplitude profiles by (i) determining the NBN frequency producing the smallest amplitude (the “dominant” frequency, of DF) and (ii) estimating 50% normalized amplitude frequencies on either side of the tip of the profile. Bandwidth (in Hz) was determined simply as the difference between the upper and lower 50% frequencies; CF was taken as the geometric mean of the upper and lower 50% frequencies. CF was also estimated from the latency-shift profiles by (i) determining the NBN frequency producing the largest latency shift (the latency DF), or (ii) from estimation of the “zero-crossing” frequency, where the latency shift went from negative (shorter) to positive (longer) – the latency CF. Narrowband masker frequencies lower than the derived-band CF tended to produce shorter latencies; masker frequencies above the CF tended to produce longer latencies. In cases where the crossing from negative-to-positive shift included a waveform with no response, a latency shift of +2.0 ms was arbitrarily assigned.^3^

### Procedure

Each subject attended two sessions, totalling about six hours in duration. After explanation of the procedures and obtaining formal consent, behavioral audiometry and determination of the broadband masking level (for subsequent HP noise level) were carried out. The order of the HP noise conditions was randomized, results for two were obtained in each session. The orders of the NBN maskers within each HP-noise condition were also randomized. All testing took place within a double-walled sound-attenuated sound booth. During testing, subjects reclined in a comfortable chair and were encouraged to sleep. Calibration of clicks, broadband noise, and NBN was checked daily.

### Results Analysis

Mean (and standard deviation) wave V %amplitude and latency shift, calculated from individual subjects relative to their individual’s nonmasked (no NBN) condition, are graphically presented for each DR condition as a function of narrowband noise masker frequency. The Dominant (DF) and Center (CF) Frequencies of the DR bands were determined from individual’s wave V %amplitude and latency-shift results; DF and CF means (and standard deviations) were assessed using repeated-measures analyses of variance (ANOVA). When significant results were found in the ANOVAs, Newman–Keuls *post hoc* tests were performed to determine the pattern of the significant differences. Results of all statistical analyses were considered significant if p<0.05.

## Results

The waveforms obtained in the NBN conditions (as well as no NBN) for the three DR frequency bands, obtained from two individual subjects and representative of the mean results, are shown in Figure 1A. Figure 1B plots the same two subjects’ results normalized (%amplitude and latency-shift) relative to their no-NBN-noise (“non-masked”) condition. The non-masked waveforms show clear waves V, with latencies increasing as the derived band frequency range decreases.^4^ Amplitudes of the non-masked (no NBN) waveforms for these two subjects are similar across the three derived bands.

**Figure 1.**
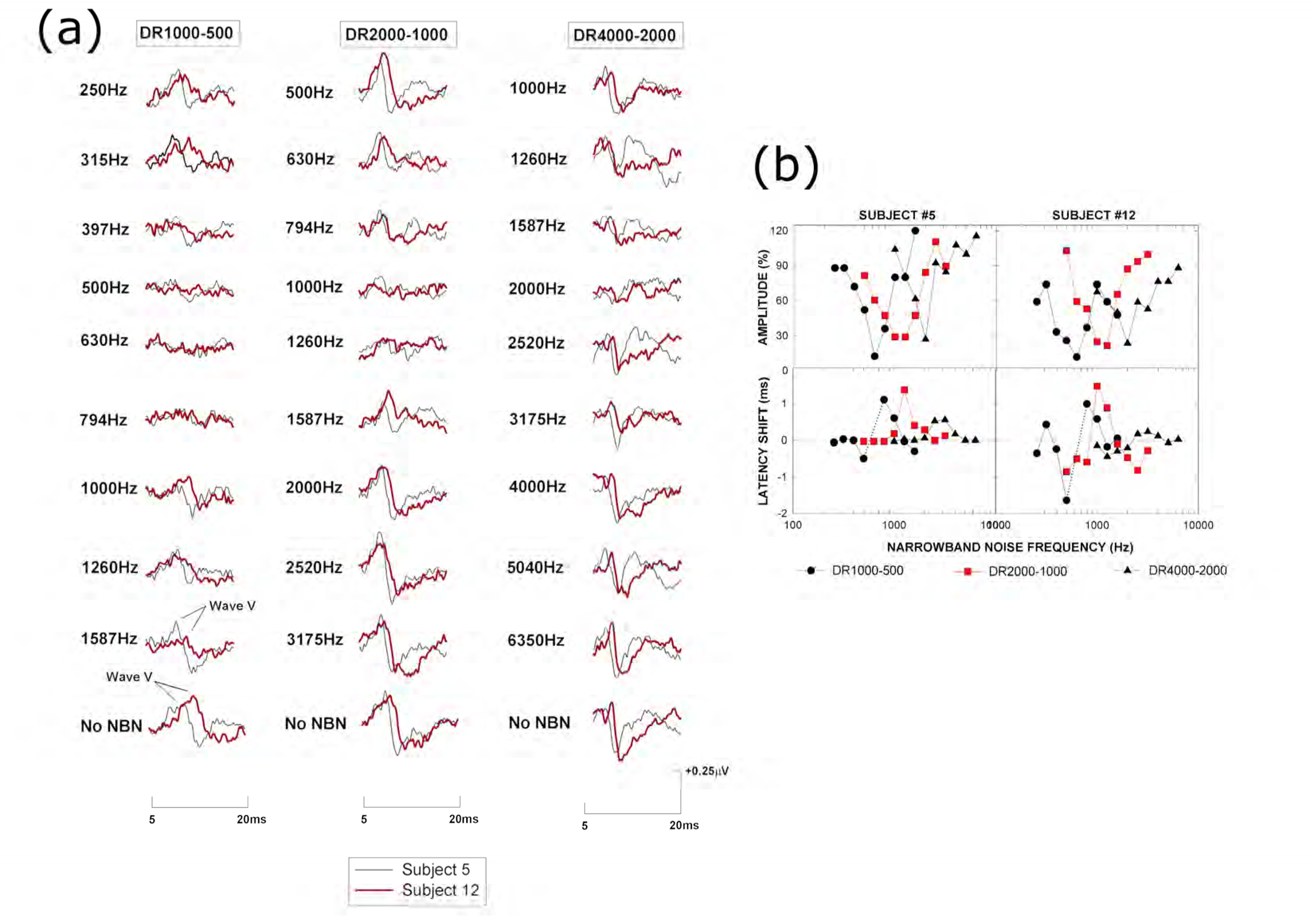
Results from two representative subjects. **(a)** Waveforms shown in each column are the narrowband noise (NBN) results for one derived band. The bottom row shows the “nonmasked” (i.e., no NBN masker) results for each derived band. **(b)** %Amplitude (top) and Latency Shift (bottom) results for Subjects 5 and 12. The dotted lines in the latency-shift graphs indicates no-response result obtained for an intermediate NBN frequency. RNL was substituted for amplitude for conditions with no response.

Although waveforms for the two subjects differ somewhat, especially in their latencies, the pattern of results with NBN maskers is essentially the same. For example: In the DR1000-500 column, there are clear waves V in the 1587-Hz down to 1000-Hz NBN masker conditions. Both subjects show no wave V in the 630-Hz NBN masker condition. Wave V clearly returns and grows in amplitude for the 397-down to 250-Hz NBN masker conditions. Thus, maximum masking occurs with the 500 to 630 Hz NBN conditions, indicating the center frequency of this derived band is likely 500 to 630 Hz. For the DR2000-1000 derived band, maximum masking for these two subjects occurs in the 1000- and 1260-Hz NBN masker conditions; for the DR4000-2000 derived band, maximum masking for these two subjects occurs in the 2000-Hz NBN masker condition. Latency-shift results are more complicated, but indicate no change (SUB #5) or shortening (SUB #12) of wave V latencies up to a NBN frequency near the DR band lower HP cutoff frequency, then an increase in latency for NBN frequencies within the DR band, with the latency shift decreasing to zero (and less) for NBN frequencies above the DR band.

Figure 2a presents the mean normalized wave V amplitudes (%Amplitude) across all subjects for each derived band, plotted as a function of NBN masker frequency. Standard deviations are plotted in the bottom panel. When a subject showed no response in a condition, the RNL for that subject’s amplitude for that condition was substituted. As these plots show clearly, each derived band is maximally masked (reduced in amplitude) by only a narrow range of frequencies, within half an octave of the lower HP frequency cutoff in the DR band. Table 1 presents the number of subjects (out of a possible 10 subjects) showing a response present in each NBN masker condition. We also calculated the %Amplitude results without the no-response RNL values, and found very similar results as are shown in Figure 2a. (Figure S1 in the Supplementary Materials section presents the results of the analysis with no-response results excluded.)

**Figure 2.**
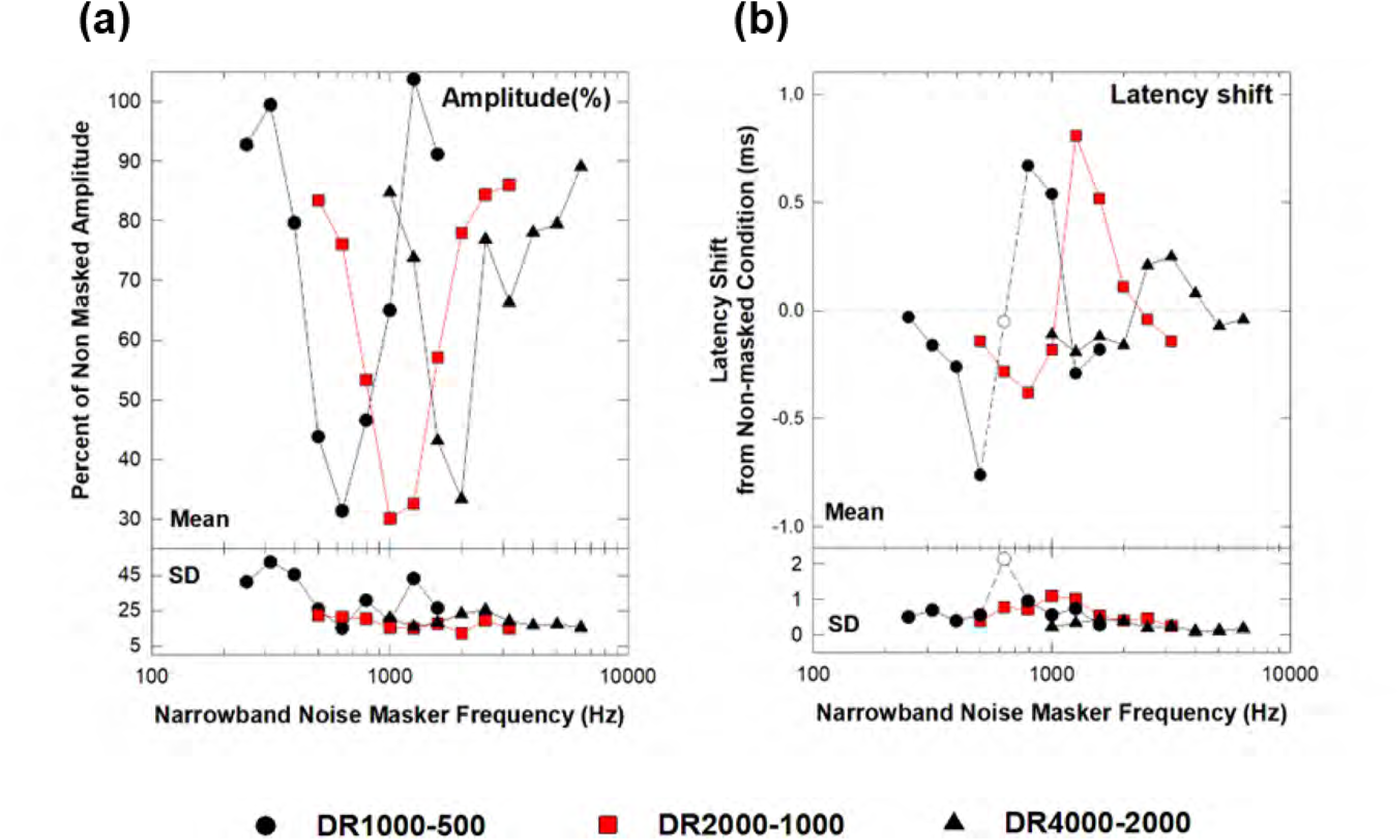
(a) Mean wave V-V’ amplitude, normalized as percent of the “nonmasked” condition (i.e., no narrowband noise), plotted as a function of NBN masker frequency. In any condition where no response was present for a subject, the RNL amplitude was inserted as amplitude. Each point represents the results from 10 subjects. SD: standard deviation. **(b)** Mean wave V latency shift, relative to the “nonmasked” (no NBN) condition, as a function of NBN masker frequency. Where a no-response result occurred, no latency shift could be determined, thus some data points are results for less than 10 subjects. Each point plotted represents the latency-shift results for at least 7 subjects, with the exception of the one point representing only 4 subjects (open circle/dashed line). Most (81%) plotted points represent data from 9-10 subjects (see Table 1). SD: standard deviation.

**Table 1.** Number of subjects with response present in each narrowband noise masker condition

|  | Narrowband Noise Masker Frequency (Hz) |  |  |  |  |  |  |  |  |
| --- | --- | --- | --- | --- | --- | --- | --- | --- | --- |
|  | 250 | 315 | 397 | 500 | 630 | 794 | 1000 | 1260 | 1587 |
| **DR1000-500** | 9 | 10 | 10 | 7 | 4 | 7 | 9 | 10 | 10 |
|  | 500 | 630 | 794 | 1000 | 1260 | 1587 | 2000 | 2520 | 3175 |
| **DR2000-1000** | 10 | 10 | 10 | 8 | 9 | 9 | 10 | 10 | 10 |
|  | 1000 | 1260 | 1587 | 2000 | 2520 | 3175 | 4000 | 5040 | 6340 |
| **DR4000-2000** | 10 | 10 | 9 | 8 | 10 | 10 | 10 | 10 | 10 |

Maximum possible = 10 subjects

Figure 2b shows the mean shift in wave V latency across all subjects, relative to the no-NBN “nonmasked” condition, as a function of NBN masker frequency. When a subject showed no response in a condition, no latency could be determined. Except for one data point (shown as an open circle), data points plotted in Figure 2b are from at least 7 subjects, with most points (81%) showing results for 9-10 subjects. The results in Figure 2b indicate the affects of NBN maskers on DR wave V latency are more complicated than those seen with amplitude: NBN maskers on the low side of the band tend to produce a *decrease* in latency; NBN maskers on the high side of the band tend to *increase* latency. This is particularly clear for the DR1000-500 results, but is also present in the results of the other two DR bands.

Each subject’s results were assessed to determine the NBN masker frequency showing either (i) the lowest amplitude or (ii) the largest latency shift, with the idea that these would indicate the dominant frequency (DF) of each DR band. Conditions with “no response” were considered smallest amplitude and largest latency shift. In a few cases, two (4/270) or three (1/270) consecutive NBN masker conditions showed lowest amplitude (or greatest latency shift); in these cases, the geometric mean of the NBN frequencies for these conditions was calculated. Table 2 presents the results of these analyses, and represents results from all 10 subjects.

**Table 2.** Mean (SD) dominant frequency (DF) of derived bands determined from minimum amplitude or maximum latency shift

|  | DR1000-500 | DR2000-1000 | DR4000-2000 |
| --- | --- | --- | --- |
| DF from minimum amplitude (Hz) | 638 (104) | 1141 (217) | 2106 (442) |
| DF from maximum latency shift (Hz) | 651 (93) | 1286 (181) | 2573 (613) |
| Number of subjects (out of 10) with DF determined by no-response results at DF | 7 | 3 | 2 |

As shown in Table 2, DR band DFs increase with the DR frequency regions, for both the %amplitude and latency-shift measures. DFs from latency-shift measures are higher than those from %amplitude measures. A 2-way repeated measures ANOVA confirms these results [Main effect for AMPLAT: F(1,9) = 9.13, p=.014]. Although differences between DF determined by latency and amplitude seem to be larger for higher DR frequencies, this did not quite reach significance [DRFREQxAMPLAT interaction: F(2,18) = 3.49; p=.053]. Expressed in octaves from the lower HP cutoffs, mean DFs determined from amplitude results are closest to the lower of the two HP cutoffs in the DR frequencies, except for DR1000-500, which is closer to 707-Hz geometric mean; DFs determined by latency are closer the geometric mean of the two HP cutoffs.

The above analysis essentially searches for the single “dominant” frequency in the NBN masker effects. However, as shown by the %amplitude and latency-shift results in Figure 2, masking effects are seen not just at one frequency but across a range of NBN frequencies. The %amplitude results were thus analyzed in order to determine (by interpolation) the frequencies of lower and upper 50% amplitude points. From these 50% cutoffs, we calculated the bandwidth (at 50% amplitude) as well as the CF (also at 50% amplitude). CF was calculated as the geometric mean of the two 50% amplitude points. Means and standard deviations of %amplitude-based results are presented in Table 3 (top).

**Table 3.**
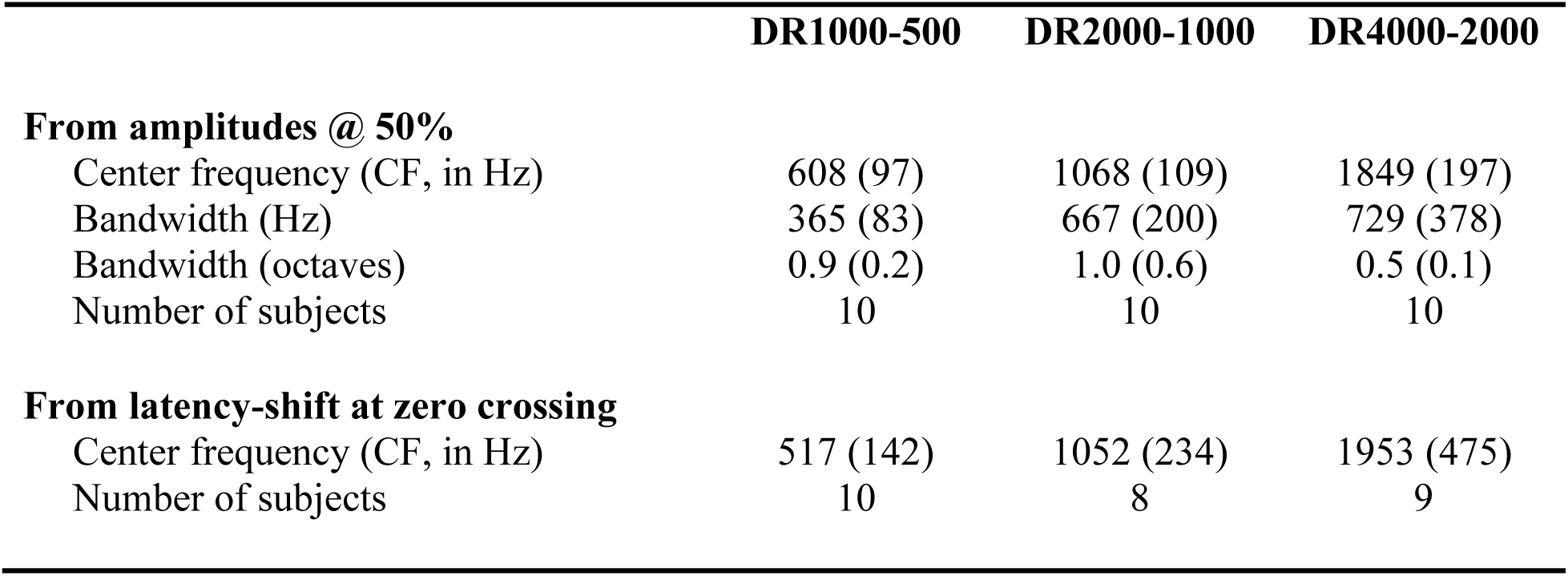
Mean (SD) of center frequencies and bandwidths of derived bands from amplitudes and latencies

|  | DR1000-500 | DR2000-1000 | DR4000-2000 |
| --- | --- | --- | --- |
| **From amplitudes @ 50%** |  |  |  |
| Center frequency (CF, in Hz) | 608 (97) | 1068 (109) | 1849 (197) |
| Bandwidth (Hz) | 365 (83) | 667 (200) | 729 (378) |
| Bandwidth (octaves) | 0.9 (0.2) | 1.0 (0.6) | 0.5 (0.1) |
| Number of subjects | 10 | 10 | 10 |
| **From latency-shift at zero crossing** |  |  |  |
| Center frequency (CF, in Hz) | 517 (142) | 1052 (234) | 1953 (475) |
| Number of subjects | 10 | 8 | 9 |

Mean DR band CFs (Table 3) derived from the %amplitude profiles are very close to the lower of the HP noise cutoff frequencies involved in each derived-band calculation, with exception of the CF for DR1000-500 which is about halfway between the lower HP noise frequency and the geometric mean of the two HP noise frequencies. (We also determined CFs from the %amplitude profiles with no-response results excluded. These results can be found in the Supplementary Materials, Table S2. CFs did not change substantially with no-response results excluded.) Table 3 also shows mean bandwidths, shown in Hz in the middle and below this in octaves, with results indicating the derived bands are narrow in frequency. The mean DR1000-500 and DR2000-1000 bandwidths, expressed as octaves, are about one octave in width. In contrast, the DR4000-2000 bandwidth was much smaller, on average only 0.5 octaves.

Determining DR band CF from the latency-shift profiles is more complicated, other than the DF analysis above that determined the NBN masker(s) that produced the largest latency shifts. However, as with the %amplitude profiles, the simple “DF” analysis fails to take into account latency effects by other NBN frequencies. The mean latency-shift results display a pattern that differs from the %amplitude results, in that both positive (longer) and negative (shorter) latency shifts were seen. As shown in Figure 2b, NBN maskers on the low side of a DR band’s frequencies tend to produce a *decrease* in latency; NBN maskers on the high side of the DR band tend to *increase* latency. Thus, going from low to high NBN masker frequency, as the noise frequency comes closer to the center frequency of the derived band, wave V latency decreases; as it goes above the CF, wave V latency increases.

Assuming the frequency region demonstrating the crossover from negative latency shift (wave V latency decrease) to positive latency shift (wave V latency increase) reflects the region of the center frequency, we therefore estimated the “zero crossing” frequency -- that is, the frequency with a shift of 0 ms -- from each subject’s latency results. Several subjects showed no response in the CF region; in these cases, in order to interpolate the “0-ms” point, we inserted +2.0 milliseconds as an estimate of the latency increase for a non-response. We chose this value based on the range of latency shifts in the data – there were several instances of such shifts (2 ms and greater) for near-threshold responses.^5^ The mean results of this latency-shift analysis are shown in Table 3 (bottom). The DR bands with fewer than 10 subjects are due to 1-2 subjects not showing the negative/positive pattern in latency shift (and hence no zero crossing could be estimated). These mean latency-shift-determined CFs, shown at the bottom of Table 3 are very similar to the CFs determined from the amplitude curves (Figure 2 and Table 3), and indeed, a 2-way repeated measures ANOVA indicates no significant difference between the CFs estimated from the %amplitude and latency-shift profiles [AMPLAT main effect: F(1,7) = 0.03, p=0.877; DRFREQ main effect: F(2,14) = 123.77, p<0.001; DRFREQxAMPLAT interaction: F(2,14) = 0.835, p=0.455]. These mean latency-shift CFs are very close to (within ±0.1 octaves) the lower HP noise cutoff. Bandwidth was not estimated from the latency-shift results due to their complexity.

## Discusssion

Overall, the results of this study confirm that DR Band ABRs, obtained using 1-octave steps between HP noise cutoffs, reflect cochlear regions representing a narrow (≤1 octave wide) range of frequencies. Using both amplitude and latency measures, the CFs of these bands are all within half an octave of the lower HP noise frequency.

The amplitude curves of the present study, in general, reflect what past studies have assumed for the DR band technique (using 1-octave HP noise steps), and lend empirical support for use of the technique. In addition to demonstrating the validity of the assumptions of the DR band technique, the results of the present study also provide further evidence concerning the CF of a DR band.

The present study analysed the results in several ways in order to determine the dominant (DF) or center (CF) frequency of each DR band. Past researchers have assumed each band’s CF to be closer to either (i) the lower HP noise cutoff frequency or (ii) the geometric mean of the band’s two HP noise cutoffs. The dominant frequencies (DF), determined from minimum amplitudes (Table 2, top), are somewhat analogous to a tuning curve’s “best” or “characteristic” frequency, and suggest the CFs for the two higher DR bands (DR4000-2000 and DR2000-1000) are very close to the lower of the two HP noise cutoffs (within 0.08-0.19 octaves), and about half way between the lower HP cutoff and the geometric mean for the for DR1000-500 (0.35 octaves above 500 Hz). The CF determined from the 50% points of the %amplitude curves (Table 3, top) showed similar results, with the two higher DR bands were closer to the lower HP noise cutoffs (−0.11 to +0.1 octaves) and the CF for DR1000-500 band being midway between the lower HP cutoff and the geometric mean (0.28 octaves from 500 Hz). On the other hand, the latency-determined DF results (Table 2, bottom) were significantly higher than the amplitude-determined DF results, with latency DFs that are a little lower than the geometric means (−-0.12 to −0.14 octaves below) for all three DR bands. CFs determined from the latency-shift zero-crossing results (Table 3, bottom), however, were all very close to the lower HP noise cutoffs (−0.03 to +0.05 octaves). Overall, all mean frequencies (CFs and DFs derived from amplitude or latency changes) were *below* the geometric mean of the two HP cutoffs in each DR band.

Although the results provide some support for both assumptions about the CF of the DR band, most measures, especially those from amplitude results, suggest the CFs are closer to the lower HP noise cutoff frequency in each DR band, with the exception of those for DR1000-500, which are about halfway between the lower HP cutoff and the geometric mean. Compared to amplitudes, latencies changes may reflect somewhat higher frequencies in the DR band, as the maximum latency (DF) results suggest the dominant frequencies are closer to the geometric mean of the two HP noise cutoffs, although this was not seen with the CFs determined from the latency shift zero-crossing. Latency changes, however, are complicated, as shown in Figure 2b. Previous researchers have also suggested that compared to amplitudes, latencies may reflect the higher frequencies contributing to responses (e.g., Burkard and Don 2007; Burkard and Hecox 1987).

In previous research using the HP/DR technique to assess place specificity of ABRs to brief tones in adults, we assumed the DR band CF was at the lower HP cutoff frequency (Nousak and Stapells 1992; Oates and Stapells 1997), and reported largest amplitudes to air- and bone-conducted tones were seen in the DR1000-500 band for 500-Hz tones and the DR4000-2000 band for 2000-Hz tones. These results are consistent with other masking studies (pure-tone or notched noise) showing best frequencies at or very close to the tone nominal frequency (e.g., Folsom and Wynne 1987; Wu and Stapells 1994). Thus, comparison of frequency-specific masking studies and DR band studies of brief-tone stimuli suggests results are more consistent with the CF of the DR band being closer to the lower HP cutoff frequency, and not the geometric mean of the two HP noise cutoffs. Additional support for the CF of DR bands being closer to the lower HP noise cutoff frequency can be found in the results of HP noise masking on a population of a cat’s auditory nerve fibres. In Evans and Elberling’s Figure 11, which shows “derived masking ratios” (a subtraction of results between adjacent HP noise cutoffs, somewhat analogous to ABR DR band), the peak masking ratio for most bands are seen at the lower HP noise cutoff involved in each band (Evans and Elberling 1982).

The bandwidth results (BW50, bandwidth at 50% amplitude) were close to one octave for the DR2000-1000 and DR1000-500 bands, as would be expected when using HP noise with 1-octave separations. The BW50 for the 4000-2000 band, however, was much smaller, on average being only 0.5 octaves (Table 3). The likely explanation for this is the rolloff of the ER3A insert earphones above 4000 Hz. Although the DR4000-2000 Hz ABR would not be reduced in amplitude by this rolloff – as shown by similar no-NBN ABR amplitudes between the DR4000-2000 and DR2000-1000 bands (Table S1 in Supplementary Materials) – the two NBN masker frequencies above 4000 Hz (5040 and 6340 Hz) would be reduced in intensity and thus less effective as maskers, resulting in a smaller BW50.

The latency-shift results show both expected and unexpected results. Increasing latency as NBN masker frequency nears the DR band center frequency would be expected. However, this effect occurs primarily for NBN masker frequencies on the high side of the DR band frequencies. On the low side of the DR band, NBN masker frequencies tended to show negative latency shifts (that is, wave V latency is shorter than that seen for the nonmasked (no NBN) DR band, especially for the DR1000-500 band (negative latency shifts were smaller for the two higher DR bands). In the present study, the latency-shift results may reflect cochlear place effect: as high-frequency contributions of a DR band are removed by NBN maskers, the remaining response shifts apically and hence is later. When low-frequency contributions are removed, wave V latency shortens as the remaining response reflects more basal contributions. The larger negative shifts seen for lower DR bands may reflect larger latency changes due to slower travelling wave velocity in apical regions. However, this pattern is not seen in the positive latency-shifts, where larger latency increases are seen in the DR2000-1000 band, next largest in the DR1000-500 band, and smallest in the DR4000-2000 band. This might be due to mechanisms in addition to cochlear place, but might be more easily explained by the greater number no-response results for the DR1000-500 band, where latencies could not be determined. Likely, however, the latency changes also reflect masking noise effects beyond changes in cochlear place, including non-cochlear-place effects, which may be due to peripheral (cochlear/VIIIth nerve) or central (brainstem and higher) effects (Burkard and Don 2007).

The amplitude and latency results differ in several ways. In contrast with the low-side negative and high-side positive latency shifts, the changes in amplitude mostly show decreases as NBN masker frequency approached the DR band frequency region, from either (frequency) side. This begs the question “Do latency and amplitude results reflect different cochlear regions within a DR band?” The analysis determining the single “dominant” frequency to a DR band (DF, Table 2) indicates latency-determined DF is significantly higher than the amplitude-determined DF, suggesting latency results reflect the higher frequencies in a DR band. This is consistent with the suggestion that wave V latencies reflect the higher cochlear frequencies contributing to a response; in other words, a basalward bias (Burkard and Don 2007; Burkard and Hecox 1987; Oates and Stapells 1997). Burkard and colleagues have shown that wave V latency remains unchanged when the bandwidth of the DR is extended to lower frequencies (with the upper HP noise cutoff held constant), indicating DR latencies primarily reflect the higher frequencies withing the DR band (Burkard and Hecox 1987). However, it must be noted that in the present study, when changes were assessed across the range of NBN masker frequencies, CFs determined from amplitude vs latency results did not differ (Table 3). All considered, the longer latencies seen with NBN maskers on the high side of the DR band frequencies is consistent with higher frequency contributions contributing to the nonmasked DR band response. However, the shortening of latency with NBN maskers on the low-frequency side of the DR band indicate that lower frequencies in the band also contribute to the latency of the nonmasked DR band wave V latency. Thus, the “highest” contributing frequencies do not solely determine wave V latency.

### Limitations of the Present Experiment

Aspects of the methodology of the present study might have resulted in the ABR better reflecting apical regions of the cochlea (compared to other HP/DR studies) but it also might have added some possible apical “bias” in the masking results. As well, the contribution of central vs peripheral masking mechanisms cannot be separated. Compared to many other HP/DR studies, this study’s use of a lower high-pass EEG filter setting (30 Hz) would have a smaller impact on contributions to the ABR from more apical cochlear regions compared to the commonly used 100-Hz HP EEG filter setting (Suzuki and Horiuchi 1977). Further, the somewhat lower stimulus intensity (50 dB nHL in this study compared 60+ dB nHL in other studies) might also reduce basalward spread of excitation. However, of greater concern, our use of NBN maskers likely produced some basal spread of masker excitation, in addition to masking the cochlear place for the NBN frequencies. Thus, some of the observed ABR changes may reflect the contributions of masking in the higher frequency regions within a given derived band. NBN maskers closer to the lower frequency region of the derived band could mask frequencies above but within the band; NBN maskers closer to the higher frequency region of the derived band would, however, be limited by the presence of the HP noise. This would result in some apical bias. In the present study, we attempted to minimize the basal spread by choosing a lower NBN masker level that only just masked (within 5 dB) the ABR in the derived band with little effect in the adjoining bands. Importantly, our finding that frequencies below the DR CF tended to shorten latencies (Figure 2b) would seem to mitigate this concern over an apical bias resulting from use of NBN maskers. Future research might investigate in more detail the effect of within-band NBN masker intensity on DR amplitudes and latencies. Some portion of the NBN (and HP noise) masking effect might also be the result of more-central masking (as opposed to cochlear place-specific effects). As shown by Burkard and colleagues, energetic masking produces greater changes with more central (wave V) compared to peripheral (wave I) waves (Burkard and Hecox 1987; Burkard and Voigt 1989; Finneran et al. 2016). A future study similar to the present study might assess DR wave I changes as a function of NBN frequency in order remove (or lessen) any contribution of central masking to these findings.

### Conclusions

This narrowband noise masking study indicates that, as generally assumed, the high-pass noise/derived response technique provides results representing frequency- (and cochlear-) specific regions, with bandwidths that are 1-octave-wide or less. The center frequencies of the derived bands are close to the lower HP cutoff frequency, except for the DR1000-500 band, which is about halfway between the lower HP cutoff and the geometric mean of the two HP cutoff frequencies. There is some evidence that latency results may reflect somewhat higher frequencies in the DR band, especially when considering the most dominant frequency (DF). These results may be specific to the parameters of this study, that is: 50-dBnHL clicks, high-pass noise with 1-octave separations between cutoffs, and a high-pass noise filter with 96 dB/octave slopes.

In addition to the general finding that the derived bands are reasonably narrow, one implication of these results is that the practice of using the “geometric mean” of the two HP cutoffs may result in small errors of approximately one-quarter to one-half an octave. This may or may not be significant, depending upon the nature of the study. For example, HP/DR studies primarily assessing latencies might show little or no error (i.e., the HP/DR CF may be close to the geometric mean) compared to studies assessing amplitudes or thresholds.

## Ethical Approval

Research Ethics Board approval was obtained from the Behavioural Research Ethics Board of the University of British Columbia prior to commencement of the study.

## Data Availability

The ABR wave V latency and amplitude data set used and/or analyzed in the current study are available from the corresponding author on reasonable request.

## Acknowledgements

Brian Schmidt provided valuable assistance with the initial pilot studies of this work. We thank Dr. Robert Burkard for his helpful feedback on drafts of this manuscript.

## SUPPLEMENTARY TABLES AND FIGURE

**Table S1.**
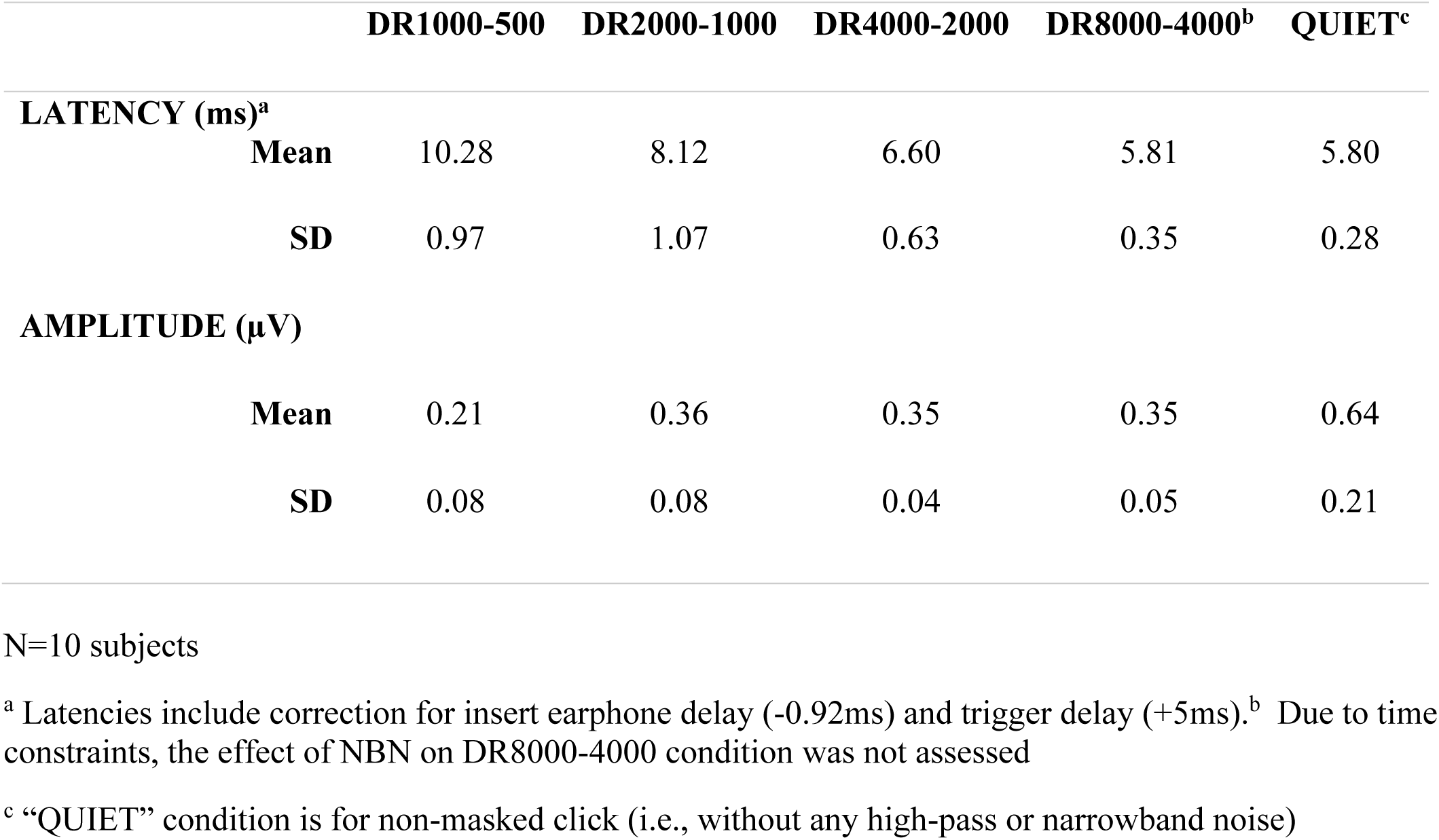
Mean wave V latencies and V-V’ amplitudes for responses *without* narrowband noise maskers.

**Table S2.**
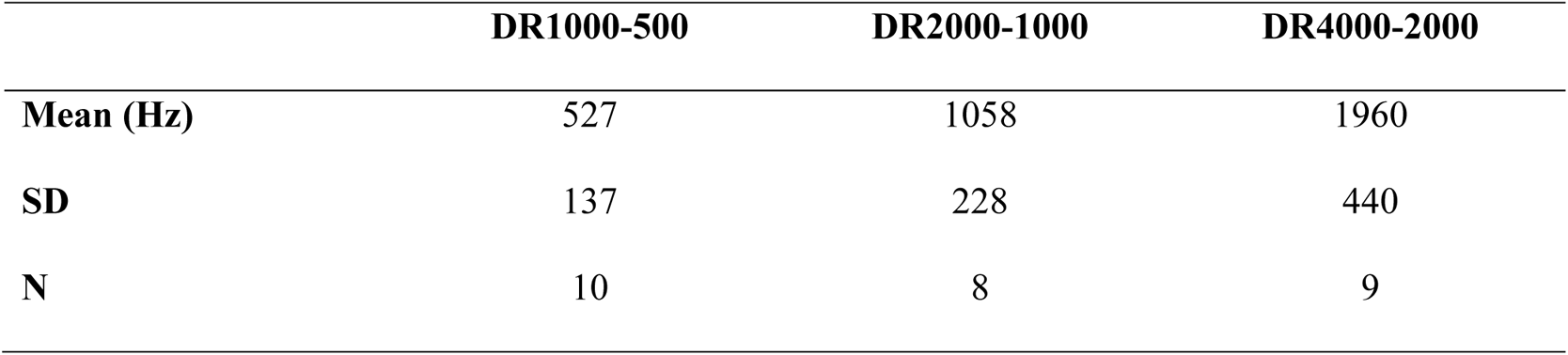
Center Frequency (CF) of the Derived Band at 50% Amplitude *with no-response results excluded*.

**Table S3.**
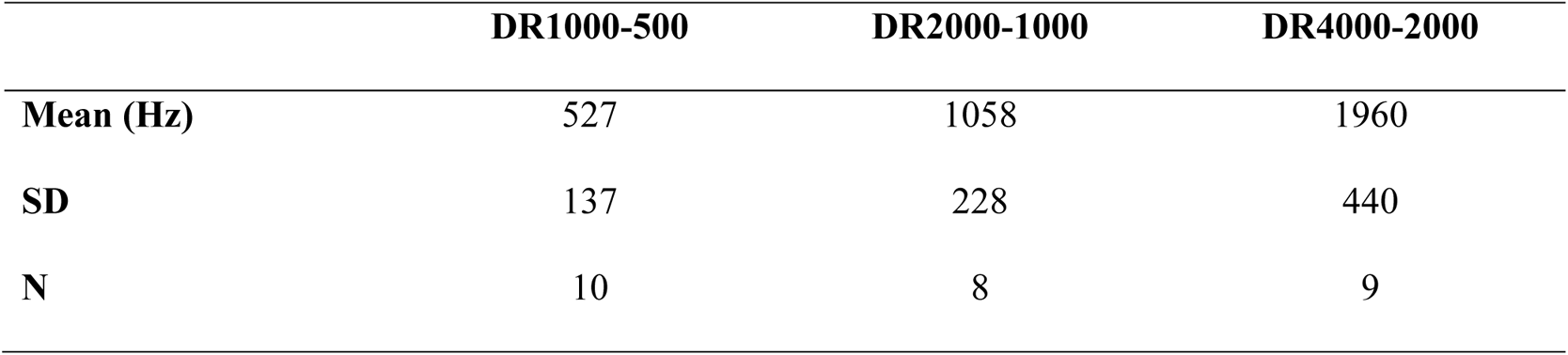
Center Frequency (CF) of the Derived Band from the latency-shift zero-crossing (using +1-ms latency shift substitution for NR)

**Figure S1.**
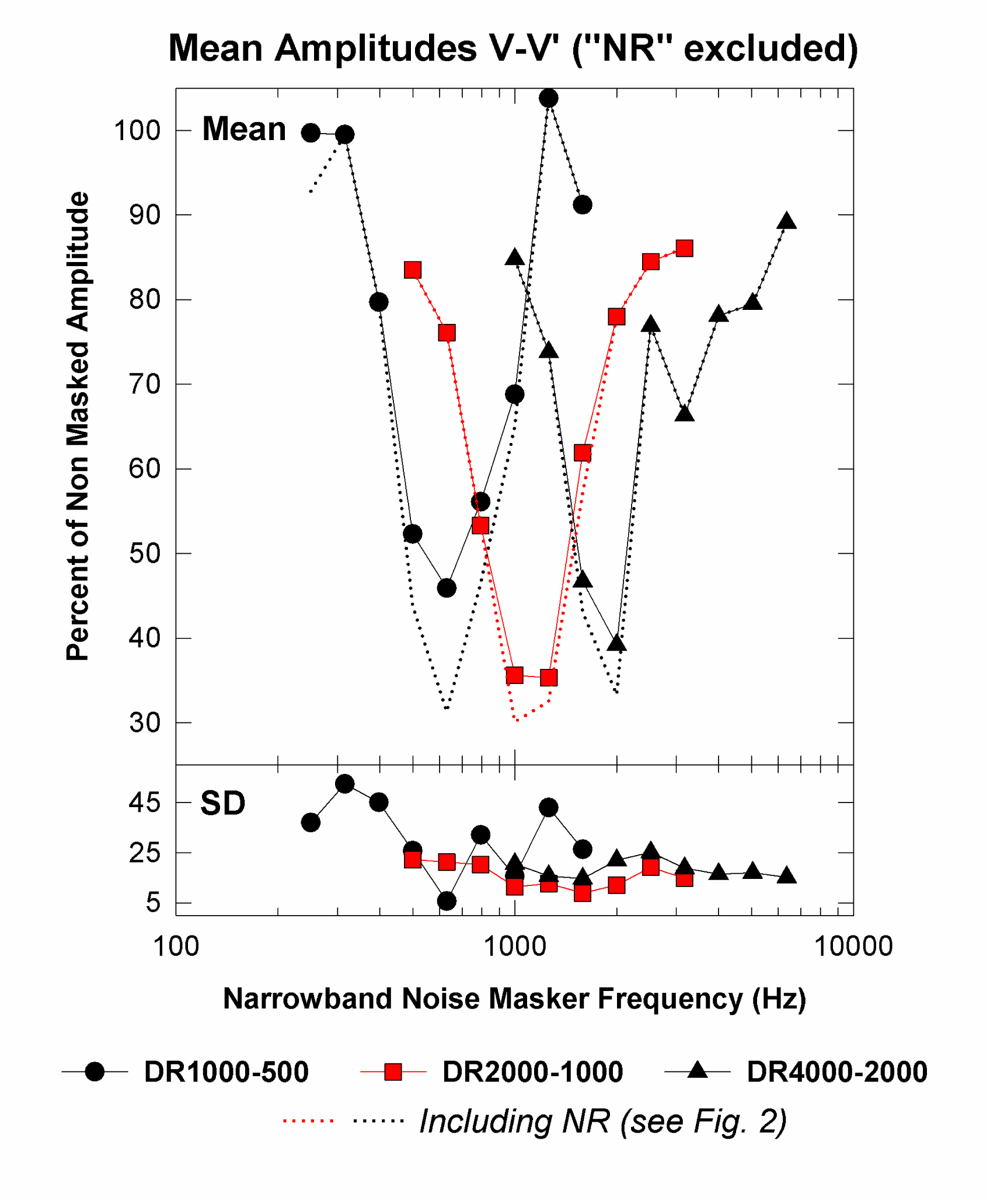
Mean and standard deviation wave V-V’ amplitude, normalized as percent of the “nonmasked” condition (i.e., no narrowband noise), plotted as a function of NBN masker frequency. No-response results removed. See Table 2 for number of subjects with response in each condition. Dotted lines represent results for all subjects including no-response results replotted from Figure 2.

## Footnotes

1 This level was determined from pilot studies to produce reduction of the derived-band ABR amplitude between the HP noise cutoffs without substantially altering the responses above or below the HP noise cutoffs (i.e., to just mask without overmasking).

2 A “no response” result indicates a substantial masking effect by the NBN masker. Thus, the amplitude for that condition would be expected to be very low; insertion of the waveform’s RNL for amplitude provided a reasonable and conservative estimate of amplitude for that condition (Herdman, Picton, and Stapells 2002). Nevertheless, Figure S1 and Table S2 in the Supplementary Materials presents the %Amplitude results with no-response results excluded; results with no-response excluded are very similar to those including no-response results shown in Figure 2 and Table 3.

3 The +2-ms latency shift for no-response results was only used to estimate the zero-crossing frequency. Mean latency-shift results shown in Figure 2b do not include this +2-ms latency shift.

4 The mean (SD) wave V latencies and V-V’ amplitudes obtained for non-masked (i.e., no NBN) derived responses are presented in the Supplementary Materials section, Table S1.

5 As a check of the +2-ms estimate, mean zero-crossing frequency results were also calculated using a smaller shift (+1 ms); the mean zero-crossing frequency results using the +1-ms shift were within 7-10 Hz of those calculated using the +2-ms shift. Table S2 in the Supplementary Materials section presents the results of the analysis using the +1-ms latency shift substitution for no response.

